# Physical Activity Decreases the Prevalence of COVID-19-associated Hospitalization: Brazil EXTRA Study

**DOI:** 10.1101/2020.10.14.20212704

**Authors:** Francis Ribeiro de Souza, Daisy Motta-Santos, Douglas dos Santos Soares, Juliana Beust de Lima, Gustavo Gonçalves Cardozo, Luciano Santos Pinto Guimarães, Carlos Eduardo Negrão, Marcelo Rodrigues dos Santos

## Abstract

**Objectives:** We compared physical activity levels before the outbreak and quarantine measures with COVID-19-associated hospitalization prevalence in surviving patients infected with SARS-CoV-2. Additionally, we investigated the association of physical activity levels with symptoms of the disease, length of hospital stay, and mechanical ventilation.

**Methods:** Between June 2020 and August 2020, we invited Brazilian survivors and fully recovered patients infected with SARS-CoV-2 to respond to an online questionnaire. We shared the electronic link to the questionnaire on the internet. In this cross-sectional study, we collected data about clinical outcomes (symptoms, medications, hospitalization, and length of hospital stay) and cofactors, such as age, sex, ethnicity, preexisting diseases, socioeconomic and educational, and physical activity levels using the International Physical Activity Questionnaire (IPAQ short version).

**Results:** Out of 938 patients, 91 (9.7%) were hospitalized due to COVID-19. In a univariate analysis, sex, age, and BMI were all associated with hospitalizations due to COVID-19. Men had a higher prevalence of hospitalization (66.6%, P=0.013). Patients older than 65 years, obese, and with preexisting disease had a higher prevalence of COVID-19-related hospitalizations. In a multivariate regression model, performance of at least 150 min/wk (moderate) and/or 75 min/wk (vigorous) physical activity was associated with a lower prevalence of hospitalizations after adjustment for age, sex, BMI, and preexisting diseases (PR=0.657; P=0.046).

**Conclusions:** Sufficient physical activity levels are associated with a lower prevalence of COVID-19-related hospitalizations. Performing at least 150 minutes a week of moderate-intensity, or 75 minutes a week of vigorous-intensity physical activity reduces this prevalence by 34.3%.

**(EXTRA SARS-CoV-2 Study, ClinicalTrials.gov number, NCT04396353)**

## INTRODUCTION

The new severe acute respiratory syndrome coronavirus 2 (SARS-CoV-2) is responsible for COVID-19 disease, initially discovered in the city of Wuhan, China, in late December 2019.^1^ In March 2020, the World Health Organization (WHO) declared SARS-CoV-2 to be a worldwide pandemic. The virus quickly spread to several continents, and strongly impacting Brazil. This pandemic claimed numerous victims with millions of confirmed cases worldwide and thousands of deaths. In Brazil alone, to date, more than 4 million cases have been confirmed with approximately 144,000 deaths, and these numbers are increasing dramatically. The main symptoms of the disease are fever, cough, shortness of breath, anosmia, and fatigue.^2^ COVID-19-associated hospitalization is an alarming issue with a higher cumulative rate in people aged 65 years and older ^3^ and with high public health care costs.

Because it is a new virus with such lethality, WHO and the governments of each country have adopted isolation and physical distancing as a preventive measure to contain the spread of the virus. So far, there are very few effective treatments, and vaccines are still being developed in ongoing clinical trials. Therefore, behavioral measures, such as personal hygiene, wearing masks, healthy nutrition, and physical activity, are potential strategies in preventing or mitigating the disease.^4 5^

It is well known that exercise training (a subtype of physical activity) improves immune system response, protecting against infections caused by intracellular microorganisms.^6^ For instance, regular exercise improves the pathogenic activity of tissue macrophages in parallel with an enhanced recirculation of immunoglobulins, anti-inflammatory cytokines, neutrophils, natural killer cells, cytotoxic T cells, and immature B cells, improving immune defense activity and healthy metabolism.^6^ Several studies suggest that being physically active could protect or minimize the effects of COVID-19 in humans.^7-9^ However, all of these studies have presented only physiological hypotheses that might explain such an association. Therefore, exercise training might be an important prevention strategy against SARS-CoV-2 and could play a key role in preventing hospitalizations and supporting faster recovery of infected patients.

The present study aimed to assess the association of physical activity before the pandemic and quarantine measures with the prevalence of hospitalizations in surviving patients infected with SARS-CoV-2 virus. Additionally, we investigated symptoms of the disease, length of hospital stay, and the use of mechanical ventilation in patients infected with SARS-CoV-2 and the association with sufficient and insufficient physical activity. We hypothesized that physically active patients diagnosed with COVID-19 have a lower number of hospitalizations compared with patients with insufficient levels of physical activity.

## METHODS

### Study design

This study is registered on ClinicalTrials.gov (NCT04396353), and its protocol has been published previously.^10^ This is a cross-sectional study comparing the physical activity levels before the pandemic and quarantine measures of patients infected with SARS-Cov-2. This study was approved by the local Human Subject Protection Committee at the Hospital das Clinicas da Faculdade de Medicina da Universidade de São Paulo (CAPPesq 5044-20-073), and all participants provided an online written informed consent. We followed the STROBE statement checklist for cross-sectional studies.^11^

Between June 2020 and August 2020, we invited Brazilian survivors and fully recovered patients infected with SARS-CoV-2 to respond to an online questionnaire. We disclosed the electronic link of the form across the country through social media, newscasts, hospitals, medical care providers, and disease control centers in some cities. The questionnaire included questions regarding clinical outcomes (symptoms, medications, hospitalization, length of hospital stay, and mechanical ventilation) and cofactors like age, sex, ethnicity, preexisting diseases, socioeconomic and educational status. The short version of the International Physical Activity Questionnaire (IPAQ)^12^ was used to collect data on physical activity levels before the outbreak and quarantine measures in surviving patients infected with SARS-CoV-2. The complete electronic questionnaire can be assessed in the supplemental material.

The inclusion criteria were men and women who had recovered and were survivors of the disease. Also included were individuals of all ages, with or without symptoms; patients with disease confirmed by a quantitative PCR viral test (qPCR), blood test (serology), and a rapid antibody test with or without hospitalization (nursery, semi-intensive and intensive unit care) and discharged from the hospital. Those with or without drug treatment, and with any chronic disease, such as diabetes, hypertension, coronary artery disease, obesity, metabolic syndrome, cancer, among others were also included. The exclusion criteria were illiterate patients who had difficulty filling out the electronic form and patients still hospitalized and with symptoms of COVID-19. In addition, after completing the form, some participants were excluded due to the quality control of the responses provided (see quality control below).

### Quality control criteria of the questionnaire responses

We collected a total of 1597 completed questionnaires, and all data were assessed to identify possible fraud and inappropriate responses. We identified 118 (7.4%) duplicate responses, which were excluded. One hundred and twenty-one (8.2%) participants were excluded because they answered that they did not have COVID-19 and did not perform any diagnostic tests. Additionally, 277 (18.7%) participants who declared that they had been infected with the SARS-CoV-2 (because they believed they were sick) but without any diagnostic test and with or without symptoms were also excluded. Finally, only 2 children under 10 years of age answered the questionnaire, and we decided to exclude them from the sample (Figure 1). After this first analysis of quality data responses, 1200 participants were selected.

**Figure 1.**
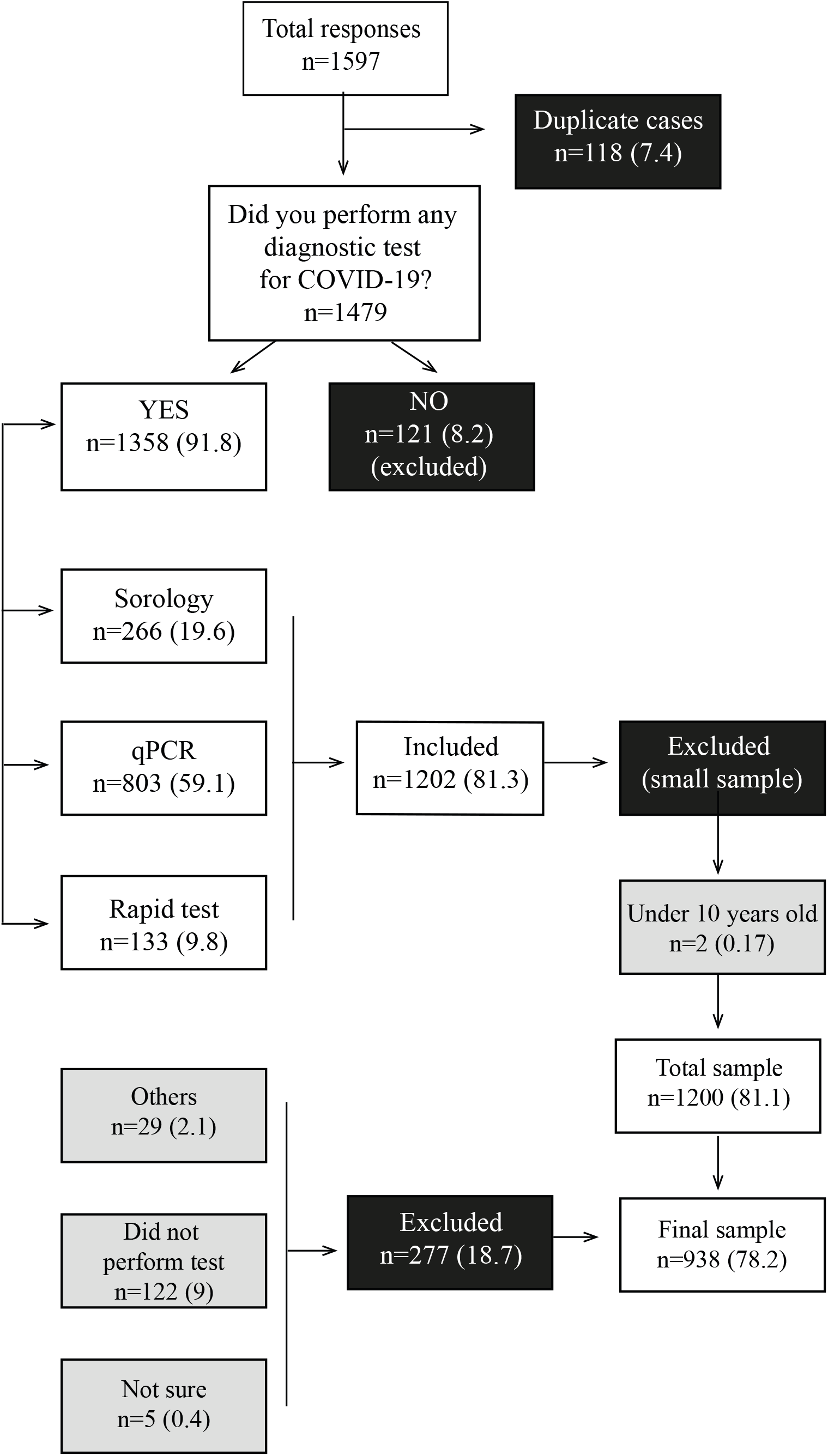
Study flowchart.

The second part of the data quality control was to correlate the self-report of physical activity with the answers provided in the IPAQ questionnaire. In one part of the questionnaire, we asked how the participants declared themselves: “athlete,” “physically active,” or “sedentary.” These 3 categories were correlated with the amount of physical activity reported in the IPAQ questionnaire. For instance, if the participant declared him/herself as sedentary, but performed more than 150 minutes of physical activity per week, he/she was excluded. After this correlation, 938 (78.2%) were included in the final analysis (Figure 1).

The participants were subdivided into 2 categories according to IPAQ information: sufficient physical activity (“athlete” and “physically active” were pooled) and insufficient physical activity (“sedentary”). Sufficient physical activity was considered when the participant performed at least 150 minutes per week of moderate physical activity and/or at least 75 minutes of vigorous physical activity.^13 14^ Insufficient physical activity was defined when the participants did not meet these recommendations. In addition, sedentary behavior was defined considering the number of hours per day spent sitting.^15^ The metabolic equivalent of task (MET) defined as the amount of oxygen consumed while sitting at rest (equal to 3.5 mL/kg/min) was calculated to estimate the energy expenditure values.^12^ Within our sample, we also calculated the 25th percentile of physical activity time (100 minutes/week) and METs (11.2 METs/hour/week); both were used to estimate the primary and secondary outcomes.

Body mass index (BMI) was categorized as follows: underweight (<18.5 kg/m^2^), normal weight (18.5-24.9 kg/m^2^), overweight (25.0-29.9 kg/m^2^), obesity I (30.0-34.9 kg/m^2^), obesity II (35.0-40.0 kg/m^2^), and obesity III (>40 kg/m^2^).

### Outcomes

The primary outcome of the study was to assess the number of hospitalizations among individuals with sufficient physical activity and insufficient physical activity who were infected with SARS-CoV-2. The secondary outcomes were to assess symptoms of the disease (such as fever, headaches and muscle aches, shortness of breath, cough, loss of taste or smell, fatigue/tiredness), length of hospital stay, and use of mechanical ventilation.

### Sample size

The sample size was calculated using the Epi-Info program, version 7.2.3.1, considering an infinite population, an unknown prevalence of 50%, a confidence level of 95%, a sampling error of 5%, and a delineation effect (deff) of 2 points. The minimum sample size was 768 subjects. Considering possible dropouts and to control for confounding factors in multivariate analysis, 10% was added resulting in a sample of 850 patients.

For the comparison of physical activity groups between the diagnoses of having or not having COVID-19, the sample was calculated based on the comparison of proportions in the G*Power software, with 80% power, 5% significance level, degree of freedom equal to 2 (following a 3×2 size table; 3=hospitalization [yes and no] and 2=physical activity [athlete, physically active, and sedentary]) and an effect size w of 0.1. The calculation of 964 subjects with the addition of 10% of losses resulted in 1061. To answer the proposed objectives, the calculation with the largest sample size was considered.

### Statistical analysis

The data analysis was performed using SPSS v.25 software. The distributions of the quantitative variables were verified by the Shapiro-Wilk normality test. Symmetric and asymmetric distribution variables are presented as mean and standard deviation or median and interquartile range (median [p25 to p75]), respectively.

Categorical variables are represented by absolute and relative frequency. The proportions of the variables were compared between the outcome (hospitalization: yes or no) using the chi-square test. When significant, post-hoc analysis was performed using Winpepi v.11.65 software.

The sample characterization variables were associated with physical activity levels by using the chi-square test. When significant, the analysis of adjusted standardized residues was used to identify local associations with values equal to or greater than 1.96.

The Poisson regression model was used to estimate crude and adjusted prevalence ratios (PR) and their respective 95% CI. The statistical significance of the PR was assessed using Wald’s statistics. Crude analysis of the variables was performed between the outcomes. After that, important multivariate outcomes were included in the model.

Among patients who were hospitalized, the association between physical activity levels and clinical variables of hospitalization (such as symptoms, medications, and mechanical ventilation) was analyzed. The distributions of length of hospital stay between physical activity levels were compared using the Mann-Whitney or Kruskal-Wallis test with the Dunn post-hoc test.

## RESULTS

Of 938 patients, 91 (9.7%) were hospitalized due to COVID-19. In a univariate analysis, sex, age, and BMI were associated with hospitalizations due to COVID-19. When degree of hospitalization was compared between men and women, men had a higher prevalence of hospitalization (66.6% [P=0.013]). Patients older than 65 years had 7 times the prevalence of hospitalization compared with adults aged 18-65 years (P<0.001). Patients with obesity I were hospitalized more than subjects with normal weight (P=0.003). When compared with eutrophic patients, those overweight and obese patients had a higher prevalence of hospitalization between 83% and 166%. Preexisting disease (P=0.002), 3 or more symptoms (P<0.001), and using 2 or more medications (P<0.001) were also factors associated with a higher prevalence of hospitalization (Table 1).

**Table 1.**
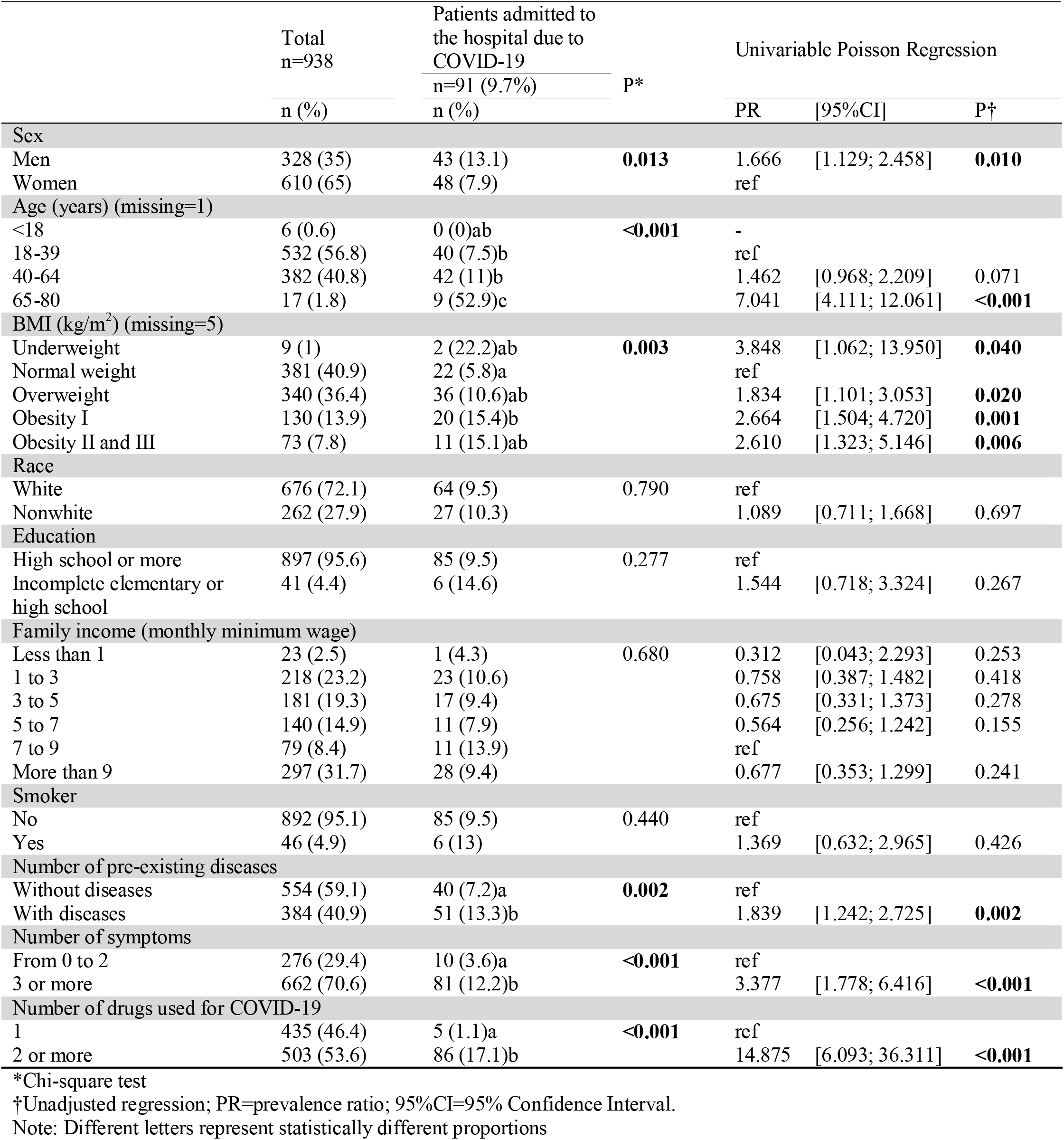
Characterization of the Sample, Comparison of Proportions and Calculation of the Crude Prevalence Ratio of the Variables in Patients With COVID-19.

All categories constructed from the IPAQ (time of physical activity in minutes per week or METs hour per week) were significant in comparison with the proportions of hospitalization (P<0.05). In a univariate analysis, patients with insufficient physical activity (mean = 62.3±86.0 min/wk and median = 25.0 min/wk [0; 106]) had a higher degree of hospitalization compared with patients with sufficient physical activity (mean = 726.9±426.7 min/wk and median = 640.0 min/wk [420; 925]). Those who were sufficiently active had a 37.6% lower prevalence of hospitalization due to COVID-19 (PR=0.624; P=0.018). Other variables of physical activity levels had a reduction in the crude prevalence of hospitalization between 35.4% (cutoff point of z11.2 METs/h/wk) and 46.2% (performs 2 or more physical activities) (Table 2).

**Table 2.** Physical Activity Level, Comparison of Proportions and Calculation of the Crude Prevalence Ratio of the Variables in Relation to Patients Hospitalized by COVID-19.

|  | Total<br>n=938 | Patients admitted<br>to the hospital due<br>to COVID-19<br>n=91 (9.7%) | P* | Univariable Poisson Regression |  |  |
| --- | --- | --- | --- | --- | --- | --- |
|  | n (%) | n (%) |  | PR | [95%CI] | P† |
| Physical activity (>150 min/wk [moderate] and/or 75 min/wk [vigorous]) |  |  |  |  |  |  |
| Sufficient | 611 (65.1) | 49 (8)a | **0.024** | 1 |  |  |
| Insufficient | 327 (34.9) | 42 (12.8)b |  | 0.624 | [0.423; 0.922] | **0.018** |
| Physical activity in minutes (25th percentile of EXTRA study) |  |  |  |  |  |  |
| ≥100 min/wk (≥6 METs/h/wk) | 707 (75.4) | 60 (8.5)a | **0.038** | ref |  |  |
| 0-99 min/wk (<6 METs/h/wk) | 231 (24.6) | 31 (13.4)b |  | 0.632 | [0.421; 0.950] | **0.027** |
| Physical activity in METs (25th percentile of EXTRA study) |  |  |  |  |  |  |
| ≥11.2 METs/h/wk | 641 (68.3) | 53 (8.3)a | **0.039** | ref |  |  |
| <11.2 METs/h/wk | 297 (31.7) | 38 (12.8)b |  | 0.646 | [0.436; 0.958] | **0.030** |
| Sitting time in hours/day (25th percentile of EXTRA study) |  |  |  |  |  |  |
| <4.7 h/day | 463 (49.4) | 51 (11) | 0.218 | ref |  |  |
| ≥4.7 h/day | 475 (50.6) | 40 (8.4) |  | 1.308 | [0.882; 1.939] | 0.181 |
| Sitting time in hours/day (75th percentile of EXTRA study) |  |  |  |  |  |  |
| <7.4 h/day | 710 (75.7) | 72 (10.1) | 0.501 | ref |  |  |
| ≥7.4 h/day | 228 (24.3) | 19 (8.3) |  | 1.217 | [0.751; 1.972] | 0.426 |
| Number of physical activities performed |  |  |  |  |  |  |
| 2 or more | 261 (27.8) | 36 (7.4)a | **0.020** | 0.538 | [0.348; 0.833] | **0.005** |
| 1 | 192 (20.5) | 19 (9.9)ab |  | 0.717 | [0.425; 1.211] | 0.214 |
| None | 485 (51.7) | 36 (13.8)b |  | ref |  |  |

\*Chi-square test
†Unadjusted regression (crude)
Note: Different letters represent statistically different proportions

Table 3 shows the characteristics between sufficient and insufficient physical activity. Men (P=0.010), normal weight (P<0.001), White race (P=0.031), with more than 9 minimum wage (monthly salary) (P<0.001), without medications (P=0.003), and without preexisting diseases (P<0.001) were all associated with sufficient physical activity. On the other hand, women, obesity I, II, and III, non-White race, less than 3 minimum wages (monthly salary), 3 or more medications, and 1 or more preexisting disease were associated with insufficient physical activity.

**Table 3.** - Proportions of Variables and Physical Activity

|  | 150 min/wk (moderate) and/or 75 min/wk (vigorous) |  | P |
| --- | --- | --- | --- |
|  | Sufficient<br>n=611 (65.1) | Insufficient<br>n=327 (34.9) |  |
|  | n (%) | n (%) |  |
| Sex |  |  |  |
| Men | 232 (38) | 96 (29.4) | 0.010 |
| Women | 379 (62) | 231 (70.6) |  |
| Age (years) |  |  |  |
| <18 | 6 (1) | 0 (0) | 0.136 |
| 18-39 | 336 (55.1) | 196 (59.9) |  |
| 40-64 | 258 (42.3) | 124 (37.9) |  |
| 65-80 | 10 (1.6) | 7 (2.1) |  |
| BMI |  |  |  |
| Under weight | 5 (0.8) | 4 (1.2) | <0.001 |
| Normal weight | 302 (49.5) | 79 (24.5) |  |
| Overweight | 234 (38.4) | 106 (32.8) |  |
| Obesity I | 58 (9.5) | 72 (22.3) |  |
| Obesity II and III | 11 (1.8) | 62 (19.2) |  |
| Race |  |  |  |
| White | 455 (74.5) | 221 (67.6) | 0.031 |
| Nonwhite | 156 (25.5) | 106 (32.4) |  |
| Education |  |  |  |
| High school or more | 509 (96.6) | 307 (93.9) | 0.081 |
| Incomplete elementary or high school | 21 (3.4) | 20 (6.1) |  |
| Family income (monthly minimum wage) |  |  |  |
| Less than 1 | 8 (1.3) | 15 (4.6) | <0.001 |
| 1 to 3 | 116 (19.0) | 102 (31.2) |  |
| 3 to 5 | 110 (18.0) | 71 (21.7) |  |
| 5 to 7 | 95 (15.5) | 45 (13.8) |  |
| 7 to 9 | 59 (9.7) | 20 (6.1) |  |
| More than 9 | 223 (36.5) | 74 (22.6) |  |
| Smoker |  |  |  |
| No | 585 (95.7) | 307 (93.9) | 0.272 |
| Yes | 26 (4.3) | 20 (6.1) |  |
| Number of drugs used for COVID-19 |  |  |  |
| None | 114 (18.7) | 40 (12.2) | 0.003 |
| 1 or 2 | 335 (54.8) | 171 (52.3) |  |
| 3 or more | 162 (26.5) | 116 (35.5) |  |
| Number of pre-existing diseases |  |  |  |
| 0 | 296 (48.4) | 139 (42.5) | <0.001 |
| 1 or more | 315 (51.6) | 188 (57.5) |  |

Chi-square test
Underlined values correspond to local association (values ≥ 1.96)

Table 4 shows the multivariate regression model to predict the factors involved in the primary outcome. Model 1 (adjusted for age and sex) showed a significant association of lower prevalence of hospitalization in patients with sufficient physical activity for the 3 categories: >150 min/wk (moderate) and/or >75 min/wk (vigorous) (PR=0.602; P=0.009), ≥100 min/wk (PR=0.666; P=0.046), and ≥11.2 METs/h/wk (PR=0.645; P=0.026). Interestingly, >150 min/wk (moderate) and/or >75 min/wk (vigorous) physical activity was independently associated with a lower prevalence of hospitalizations in model 2 (adjusted for age, sex, and BMI, PR=0.642; P=0.028) and model 3 (adjusted for age, sex, and preexisting diseases, PR=0.657; P=0.046).

**Table 4.** Poisson Multivariate Regression Model.

|  | Model 1 |  |  | Model 2 |  |  | Model 3 |  |  |
| --- | --- | --- | --- | --- | --- | --- | --- | --- | --- |
|  | PR | [CI 95%] | P | PR | [CI 95%] | P | PR | [CI 95%] | P |
| Physical activity (>150 min/wk [moderate] and/or 75 min/wk [vigorous]) |  |  |  |  |  |  |  |  |  |
| Sufficient | 0.602 | [0.411; 0.883] | **0.009** | 0.642 | [0.432; 0.954] | **0.028** | 0.657 | [0.435; 0.992] | **0.046** |
| Insufficient | 1 |  |  | 1 |  |  | 1 |  |  |
| Physical activity in minutes (25th percentile of EXTRA study) |  |  |  |  |  |  |  |  |  |
| ≥100 min/wk | 0.666 | [0.446; 0.993] | **0.046** | 0.710 | [0.470; 1.074] | 0.105 | 0.729 | [0.476; 1.115] | 0.145 |
| 0-99 min/wk | 1 |  |  | 1 |  |  | 1 |  |  |
| Physical activity in METs (25th percentile of EXTRA study) |  |  |  |  |  |  |  |  |  |
| ≥11.2 METs/h/wk | 0.645 | [0.439; 0.949] | **0.026** | 0.687 | [0.461; 1.022] | 0.064 | 0.708 | [0.467; 1.073] | 0.104 |
| <11.2 METs/h/wk | 1 |  |  | 1 |  |  | 1 |  |  |
| Sitting time in hours/day (25th percentile of EXTRA study) |  |  |  |  |  |  |  |  |  |
| <4.7 | 1.179 | [0.798; 1.743] | 0.408 | 1.266 | [0.851; 1.883] | 0.244 | 1.241 | [0.836; 1.841] | 0.284 |
| ≥4.7 | 1 |  |  | 1 |  |  | 1 |  |  |
| Sitting time in hours/day (75th percentile of EXTRA study) |  |  |  |  |  |  |  |  |  |
| <7.4 | 1.092 | [0.679; 1.758] | 0.716 | 1.133 | [0.704; 1.822] | 0.608 | 1.141 | [0.709; 1.838] | 0.586 |
| ≥7.4 | 1 |  |  | 1 |  |  | 1 |  |  |

Model 1: adjusted for age (continuous) and sex
Model 2: adjusted for age (continuous), sex, and BMI (continuous)
Model 3: adjusted for age (continuous), sex, and pre-existing diseases (yes or no)

Tables S5 and S6 (supplemental materials) show the comparison among hospitalized patients regarding intubation, oxygen therapy, symptoms, and medications used. Notably, there were no significant differences between these clinical outcomes and physical activity levels.

## DISCUSSION

The present study aimed to assess the association of physical activity before the pandemic and quarantine measures with the prevalence of hospitalizations in surviving patients infected with SARS-CoV-2 virus. The main finding is that sufficient physical activity as recommended by WHO^14 16^ decreases the prevalence of COVID-19-related hospitalization. Interestingly, performing at least 150 minutes a week of moderate-intensity, or 75 minutes a week of vigorous-intensity aerobic physical activity, or an equivalent combination of moderate- and vigorous-intensity aerobic activity, is associated with a 34.3% lower prevalence of hospitalization due to COVID-19 (PR=0.657; P=0.046). However, physical activity has no additional protection among those patients who were hospitalized. There was no difference in the prevalence of intubation, oxygen therapy, symptoms, and length of hospital stay between physically active and inactive patients.

To date, Brazil has experienced mortality of approximately 3% (68.72 deaths/100,000 population^17^), and COVID-19 has been associated with a higher incidence of hospitalizations ^18^. Approximately 8% of confirmed cases (more than 389,000 patients) required hospital care. This burden has a high public health impact with an estimated cost of US$13,000 for less severe patients and US$40,000 for more severe patients with ventilator support.^19^ Several studies have shown that obesity, preexisting diseases, and advanced age are important predictors of the severity of COVID-19.^2 20 21^ Our study extends the knowledge that physical activity is a marker of hospitalization prediction after adjusting for these cofactors.

In the present study, we observed the higher prevalence of hospitalization in older patients, which could be explained by the association of systemic inflammation (termed “inflammaging^22^”) in elderly people. Interestingly, exercise training attenuates aging biomarkers and modulates the redox balance.^19^ Master athletes have better redox balance and inflammatory status compared with middle age-matched controls.^23^ Thus, physical activity may be a strategy to prevent clinical severity in elderly persons with COVID-19.

Obesity has been associated with insufficient physical activity and sedentary behavior.^24-27^ Indeed, we found a higher proportion of obese patients with insufficient physical activity. Inflammation has been described as a key mechanism in the worsening of COVID-19 in the obese.^28 29^ Interestingly, physical activity is recommended as part of weight loss for obese adults (Evidence Category A).^25 30 31^ In addition, physical activity is one of the nonpharmacological therapies to reduce inflammatory cytokines,^32^ which may be a possible mechanism of protection in patients infected with SARS-CoV-2. It has been suggested that central obesity compromises lung ventilation and may exacerbate COVID-19.^33^ Increased abdominal obesity^34^ and obstructive sleep apnea^35^ are common conditions in obesity, and both are remarkably improved after an exercise training program.^25 30^

Physical activity improves the immune system response mainly due to the amelioration of recirculation of immunoglobulins, neutrophils, natural killer cells, cytotoxic T cells, and immature B cells.^6^ Additionally, exercise prevents and treats numerous COVID-19-associated complications, such as cardiac, neurological, and metabolic disorders,^27 36^ including the positive effect on the renin-angiotensin system.^37-39^ Future studies are needed to better understand the mechanisms involved in physical activity and its benefits in patients with COVID-19.

## STUDY LIMITATIONS

This is an observational study and the results should be interpreted with caution. Although we found an association between physical activity and lower prevalence of COVID-19-related hospitalizations, we did not investigate the mechanisms involved in this relationship. All participants filled out the electronic form by themselves and possible bias should be considered. However, we constructed a quality control criterion to reduce such biases. We did not directly measure physical activity by an accelerometer device; however, self-report physical activity questionnaires have been largely used in research, and a systematic review shows that the IPAQ short form has excellent test–retest reliability (r=0.91).^40^

## CONCLUSION

Sufficient physical activity is associated with a lower prevalence of COVID-19-related hospitalizations. Performing at least 150 minutes a week of moderate-intensity, or 75 minutes a week of vigorous-intensity physical activity reduces this prevalence by 34.3%. Public policies should encourage regular physical exercise to prevent complications from COVID-19.

## Supporting information

Portuguese Google Form

English Google Form

## Data Availability

All data are shared publicly through the tool Open Science Framework (https://osf.io/crv6t).

## WHAT ARE THE NEW FINDINGS?

- Performing at least 150 minutes a week of moderate-intensity, or 75 minutes a week of vigorous-intensity aerobic physical activity, or an equivalent combination of moderate- and vigorous-intensity aerobic activity, is associated with a 34.3% lower prevalence of hospitalization due to COVID-19.

## HOW MIGHT IT IMPACT ON CLINICAL PRACTICE IN THE FUTURE?

- To date, there are very few effective treatments against SARS-CoV-2 and vaccines are still being developed in ongoing clinical trials.
- Physical activity is an important prevention strategy against SARS-CoV-2 and can play a key role in preventing hospitalizations. Public policies should encourage regular physical exercise to prevent complications from COVID-19.

## PATIENT AND PUBLIC INVOLVEMENT

Patients and/or the public were not involved in the design, or conduct, or reporting, or dissemination plans of this research.

## DATA SHARING

All data are shared publicly through the tool Open Science Framework (https://osf.io/crv6t).

## CONTRIBUTIONS

Conception of the study: F.R.S., D.M.S., D.S.S., J.B.L., G.G.C., C.E.N., and M.R.S.

Major draft of the protocol: F.R.S., D.M.S., D.S.S., J.B.L., G.G.C., C.E.N., and M.R.S.

Minor draft of the protocol: L.S.P.G.

Statistical analysis: L.S.P.G.

## REFERENCES

1. Zhu N, Zhang D, Wang W, et al. A Novel Coronavirus from Patients with Pneumonia in China, 2019. N Engl J Med 2020;382(8):727–33. doi: 10.1056/NEJMoa2001017 [published Online First: 2020/01/24]

2. Richardson S, Hirsch JS, Narasimhan M, et al. Presenting Characteristics, Comorbidities, and Outcomes Among 5700 Patients Hospitalized With COVID-19 in the New York City Area. JAMA 2020;323(20):2052–59. doi: 10.1001/jama.2020.6775

3. Du RH, Liang LR, Yang CQ, et al. Predictors of mortality for patients with COVID-19 pneumonia caused by SARS-CoV-2: a prospective cohort study. Eur Respir J 2020;55(5) doi: 10.1183/13993003.00524-2020 [published Online First: 2020/05/07]

4. World Health O. Advice on the use of masks in the context of COVID-19: interim guidance, 5 June 2020. Geneva: World Health Organization, 2020.

5. World Health O. Infection prevention and control during health care when coronavirus disease (COVID-19) is suspected or confirmed: interim guidance, 29 June 2020. >Geneva: World Health Organization, 2020.

6. Nieman DC, Wentz LM. The compelling link between physical activity and the body’s defense system. J Sport Health Sci 2019;8(3):201–17. doi: 10.1016/j.jshs.2018.09.009 [published Online First: 2018/11/16]

7. Zbinden-Foncea H, Francaux M, Deldicque L, et al. Does High Cardiorespiratory Fitness Confer Some Protection Against Proinflammatory Responses After Infection by SARS-CoV-2? Obesity (Silver Spring) 2020 doi: 10.1002/oby.22849 [published Online First: 2020/04/23]

8. Dixit S. Can moderate intensity aerobic exercise be an effective and valuable therapy in preventing and controlling the pandemic of COVID-19? Med Hypotheses 2020;143:109854. doi: 10.1016/j.mehy.2020.109854 [published Online First: 2020/05/20]

9. Amatriain-Fernández S, Gronwald T, Murillo-Rodríguez E, et al. Physical Exercise Potentials Against Viral Diseases Like COVID-19 in the Elderly. Front Med (Lausanne) 2020;7:379. doi: 10.3389/fmed.2020.00379 [published Online First: 2020/07/03]

10. Souza FRD, Motta-Santos D, Soares DDS, et al. The Impact of EXercise TRAining, Physical Activity and Sedentary Lifestyle on Clinical Outcomes in Surviving Patients Infected with the SARS-CoV-2 Virus (EXTRA SARS-CoV-2 Study): a Protocol. Web: OSF, 2020.

11. Vandenbroucke JP, von Elm E, Altman DG, et al. Strengthening the Reporting of Observational Studies in Epidemiology (STROBE): explanation and elaboration. Int J Surg 2014;12(12):1500–24. doi: 10.1016/j.ijsu.2014.07.014 [published Online First: 2014/07/18]

12. Craig CL, Marshall AL, Sjöström M, et al. International physical activity questionnaire: 12-country reliability and validity. Med Sci Sports Exerc 2003;35(8):1381–95. doi: 10.1249/01.MSS.0000078924.61453.FB

13. Guthold R, Stevens GA, Riley LM, et al. Worldwide trends in insufficient physical activity from 2001 to 2016: a pooled analysis of 358 population-based surveys with 1·9 million participants. Lancet Glob Health 2018;6(10):e1077–e86. doi: 10.1016/S2214-109X(18)30357-7 [published Online First: 2018/09/04]

14. Piercy KL, Troiano RP, Ballard RM, et al. The Physical Activity Guidelines for Americans. JAMA 2018;320(19):2020–28. doi: 10.1001/jama.2018.14854

15. Atkin AJ, Gorely T, Clemes SA, et al. Methods of Measurement in epidemiology: sedentary Behaviour. Int J Epidemiol 2012;41(5):1460–71. doi: 10.1093/ije/dys118

16. World Health O. Global recommendations on physical activity for health. Geneve, 2010.

17. Center TJHCR. Mortality Analysis 2020 [Available from: https://coronavirus.jhu.edu/data/mortality accessed September 17, 2020 at 03:00 AM EDT 2020.

18. Brasil MdS. Boletim Epidemiológico Especial: Doença pelo Coronavírus COVID-19. Semana Epidemiológica 38(13 a 19/09) de 2020. Brasília/DF, 2020.

19. Levitt L, Schwartz K, Lopez EL. Estimated Cost of Treating the Uninsured Hospitalized with COVID-19 Web2020 [Available from: https://www.kff.org/coronavirus-covid-19/issue-brief/estimated-cost-of-treating-the-uninsured-hospitalized-with-covid-19/ accessed September 17 2020.

20. Goyal P, Choi JJ, Pinheiro LC, et al. Clinical Characteristics of Covid-19 in New York City. N Engl J Med 2020;382(24):2372–74. doi: 10.1056/NEJMc2010419 [published Online First: 2020/04/17]

21. Zhou F, Yu T, Du R, et al. Clinical course and risk factors for mortality of adult inpatients with COVID-19 in Wuhan, China: a retrospective cohort study. Lancet 2020;395(10229):1054–62. doi: 10.1016/S0140-6736(20)30566-3 [published Online First: 2020/03/11]

22. Franceschi C, Campisi J. Chronic inflammation (inflammaging) and its potential contribution to age-associated diseases. J Gerontol A Biol Sci Med Sci 2014;69 Suppl 1:S4–9. doi: 10.1093/gerona/glu057

23. Rosa TS, Neves RVP, Deus LA, et al. Sprint and endurance training in relation to redox balance, inflammatory status and biomarkers of aging in master athletes. Nitric Oxide 2020;102:42–51. doi: 10.1016/j.niox.2020.05.004 [published Online First: 2020/06/18]

24. Heymsfield SB, Wadden TA. Mechanisms, Pathophysiology, and Management of Obesity. N Engl J Med 2017;376(15):1492. doi: 10.1056/NEJMc1701944

25. Villareal DT, Aguirre L, Gurney AB, et al. Aerobic or Resistance Exercise, or Both, in Dieting Obese Older Adults. N Engl J Med 2017;376(20):1943–55. doi: 10.1056/NEJMoa1616338

26. Biddle SJH, García Bengoechea E, Pedisic Z, et al. Screen Time, Other Sedentary Behaviours, and Obesity Risk in Adults: A Review of Reviews. Curr Obes Rep 2017;6(2):134–47. doi: 10.1007/s13679-017-0256-9

27. Booth FW, Roberts CK, Thyfault JP, et al. Role of Inactivity in Chronic Diseases: Evolutionary Insight and Pathophysiological Mechanisms. Physiol Rev 2017;97(4):1351–402. doi: 10.1152/physrev.00019.2016

28. Kassir R. Risk of COVID-19 for patients with obesity. Obes Rev 2020 doi: 10.1111/obr.13034 [published Online First: 2020/04/13]

29. Mauvais-Jarvis F. Aging, Male Sex, Obesity, and Metabolic Inflammation Create the Perfect Storm for COVID-19. Diabetes 2020;69(9):1857–63. doi: 10.2337/dbi19-0023 [published Online First: 2020/07/15]

30. Villareal DT, Chode S, Parimi N, et al. Weight loss, exercise, or both and physical function in obese older adults. N Engl J Med 2011;364(13):1218–29. doi: 10.1056/NEJMoa1008234

31. Donnelly JE, Blair SN, Jakicic JM, et al. American College of Sports Medicine Position Stand. Appropriate physical activity intervention strategies for weight loss and prevention of weight regain for adults. Med Sci Sports Exerc 2009;41(2):459–71. doi: 10.1249/MSS.0b013e3181949333

32. Gleeson M, Bishop NC, Stensel DJ, et al. The anti-inflammatory effects of exercise: mechanisms and implications for the prevention and treatment of disease. Nat Rev Immunol 2011;11(9):607–15. doi: 10.1038/nri3041

33. Franssen FM, O’Donnell DE, Goossens GH, et al. Obesity and the lung: 5. Obesity and COPD. Thorax 2008;63(12):1110–7. doi: 10.1136/thx.2007.086827

34. Sanchis-Gomar F, Lavie CJ, Mehra MR, et al. Obesity and Outcomes in COVID-19: When an Epidemic and Pandemic Collide. Mayo Clin Proc 2020;95(7):1445–53. doi: 10.1016/j.mayocp.2020.05.006 [published Online First: 2020/05/19]

35. Maki-Nunes C, Toschi-Dias E, Cepeda FX, et al. Diet and exercise improve chemoreflex sensitivity in patients with metabolic syndrome and obstructive sleep apnea. Obesity (Silver Spring) 2015;23(8):1582–90. doi: 10.1002/oby.21126 [published Online First: 2015/07/06]

36. Kujala UM. Evidence on the effects of exercise therapy in the treatment of chronic disease. Br J Sports Med 2009;43(8):550–5. doi: 10.1136/bjsm.2009.059808 [published Online First: 2009/04/29]

37. Negrão CE, Middlekauff HR. Exercise training in heart failure: reduction in angiotensin II, sympathetic nerve activity, and baroreflex control. J Appl Physiol 2008;104(3):577–8. doi: 10.1152/japplphysiol.01368.2007

38. Frantz EDC, Prodel E, Braz ID, et al. Modulation of the renin-angiotensin system in white adipose tissue and skeletal muscle: focus on exercise training. Clin Sci (Lond) 2018;132(14):1487–507. doi: 10.1042/CS20180276 [published Online First: 2018/07/23]

39. Rush JW, Aultman CD. Vascular biology of angiotensin and the impact of physical activity. Appl Physiol Nutr Metab 2008;33(1):162–72. doi: 10.1139/H07-147

40. Silsbury Z, Goldsmith R, Rushton A. Systematic review of the measurement properties of self-report physical activity questionnaires in healthy adult populations. BMJ Open 2015;5(9):e008430. doi: 10.1136/bmjopen-2015-008430 [published Online First: 2015/09/15]

