## Supplementary material for "Physical Activity Decreases the Prevalence of COVID-19-associated Hospitalization: Brazil EXTRA Study": Tables S5 and S6 (supplemental materials)

**Table S5** – Intubation and Oxygen Therapy Among Hospitalized Patients and Physical Activity

|  | Patients hospitalized | | | | |  | Length of hospital stay†(n=87) | |
| --- | --- | --- | --- | --- | --- | --- | --- | --- |
|  | Intubation  (n=91) |  | Oxygen therapy  (n=91) |  | Spontaneous breathing  (n=91) |  |  |  |
|  | n (%) |  | n (%) |  | n (%) |  | Median [P25; P75] | n |
| Physical activity (>150 min/wk [moderate] and/or 75 min/wk [vigorous]) | | | | | | | | |
| Sufficient | 3/49 (6.1) |  | 25/49 (51) |  | 23/49 (46.9) |  | 6.00 [4.00; 9.00] | 49 |
| Insufficient | 6/42 (14.3) |  | 20/42 (47.6) |  | 20/42 (47.6) |  | 6.00 [3.00; 12.00] | 38 |
| P | 0.293 |  | 0.910 |  | >0.999 |  | 0.874* | |
| Physical activity in minutes (25th percentile of EXTRA study) | | | | | | | | |
| ≥100 min/wk (≥6 METs/h/wk) | 3/60 (5) |  | 30/60 (50) |  | 30/60 (50) |  | 5.50 [4.00; 8.75] | 60 |
| 0-99 min/wk (<6 METs/h/wk) | 6/31 (19.4) |  | 15/31 (48.4) |  | 13/31 (41.9) |  | 6.00 [3.00; 14.00] | 27 |
| P | 0.058 |  | >0.999 |  | 0.611 |  | 0.244* | |
| Physical activity in METs (25th percentile of EXTRA study) | | | | | | | | |
| ≥11.2 METs/h/wk | 3/53 (5.7) |  | 27/53 (50.9) |  | 26/53 (49.1) |  | 6.00 [4.00; 8.50] | 53 |
| <11.2 METs/h/wk | 6/38 (15.8) |  | 18/38 (47.4) |  | 17/38 (44.7) |  | 6.00 [3.00; 12.50] | 34 |
| P | 0.112 |  | 0.901 |  | 0.846 |  | 0.404* | |
| Sitting time in hours/day (25th percentile of EXTRA study) | | | | | | | | |
| <4.7 h/day | 6/51 (11.8) |  | 29/51 (56.9) |  | 22/51 (43.1) |  | 6.00 [4.00; 9.50] | 49 |
| ≥4.7 h/day | 3/40 (7.5) |  | 16/40 (40) |  | 21/40 (52.5) |  | 5.00 [3.75; 10.00] | 38 |
| P | 0.726 |  | 0.166 |  | 0.499 |  | 0.424* | |
| Sitting time in hours/day (75th percentile of EXTRA study) | | | | | | | | |
| <7.4 | 7/72 (9.7) |  | 37/72 (51.4) |  | 33/72 (45.8) |  | 6.00 [4.00; 10.00] | 70 |
| ≥7.4 | 2/19 (10.5) |  | 8/19 (42.1) |  | 10/19 (52.6) |  | 5.00 [3.50; 7.50] | 17 |
| P | >0.999 |  | 0.644 |  | 0.787 |  | 0.381* | |
| Chi-square test | | | | | | | | |
| *Mann-Whitney test  †variable with missing value | | | | | | | | |

**Table S6** - Comparison of the proportion of symptoms and medication used among hospitalized patients (n = 91)

|  | 150 min/wk (moderate) and/or 75 min/wk (vigourous) | | |  |  |  |  | 150 min/wk (moderate) and/or 75 min/wk (vigourous) | | |  |  |
| --- | --- | --- | --- | --- | --- | --- | --- | --- | --- | --- | --- | --- |
|  | Insufficient |  | Sufficient |  | P |  |  | Insufficient |  | Sufficient |  | P |
|  | n (%) |  | n (%) |  |  |  |  | n (%) |  | n (%) |  |  |
| Body pain | | | | | |  | Antibiotics | | | | | |
| No | 8 (16.3) |  | 9 (21.4) |  | 0.724 |  | No | 5 (10.2) |  | 2 (4.8) |  | 0.445 |
| Yes | 41 (83.7) |  | 33 (78.6) |  |  |  | Yes | 44 (89.8) |  | 40 (95.2) |  |  |
| Fatigue/tiredness | | | | | |  | Anticoagulant | | | | | |
| No | 7 (14.3) |  | 7 (16.7) |  | 0.982 |  | No | 15 (30.6) |  | 11 (26.2) |  | 0.816 |
| Yes | 42 (85.7) |  | 35 (83.3) |  |  |  | Yes | 34 (69.4) |  | 31 (73.8) |  |  |
| Shortness of breath | | | | | |  | Antiviral | | | | | |
| No | 12 (24.5) |  | 14 (33.3) |  | 0.485 |  | No | 31 (63.3) |  | 35 (83.3) |  | 0.057 |
| Yes | 37 (75.5) |  | 28 (66.7) |  |  |  | Yes | 18 (36.7) |  | 7 (16.7) |  |  |
| Fever | | | | | |  | Corticosteroids | | | | | |
| No | 49 (100) |  | 40 (95.2) |  | 0.210 |  | No | 26 (53.1) |  | 25 (59.5) |  | 0.684 |
| Yes | 0 (0) |  | 2 (4.8) |  |  |  | Yes | 23 (46.9) |  | 17 (40.5) |  |  |
| Loss of taste or smell | | | | | |  | Hydroxychloroquine or chloroquine | | | | | |
| No | 14 (28.6) |  | 16 (38.1) |  | 0.459 |  | No | 32 (65.3) |  | 28 (66.7) |  | >0.999 |
| Yes | 35 (71.4) |  | 26 (61.9) |  |  |  | Yes | 17 (34.7) |  | 14 (33.3) |  |  |
| Cough | | | | | |  | For pain or fever | | | | | |
| No | 10 (20.4) |  | 14 (33.3) |  | 0.233 |  | No | 49 (100) |  | 42 (100) |  | - |
| Yes | 39 (79.6) |  | 28 (66.7) |  |  |  | Yes | - |  | - |  |  |
| *Chi-square test | | | | | | | | | | | | |
