## Supplementary material for "Physical Activity Decreases the Prevalence of COVID-19-associated Hospitalization: Brazil EXTRA Study": Portuguese Google Form

### Título da pesquisa: O Impacto do Exercício Físico, da Atividade Física e do Sedentarismo nos Desfechos Clínicos em Pacientes Sobreviventes Infectados pelo Vírus SARS-CoV-2 (Coronavírus)

HOSPITAL DAS CLÍNICAS DA FACULDADE DE MEDICINA DA UNIVERSIDADE DE SÃO PAULO-HCFMUSP

TERMO DE CONSENTIMENTO LIVRE E ESCLARECIDO (TCLE)

Esse formulário será utilizado para um estudo científico do Instituto do Coração do Hospital das Clínicas da Universidade de São Paulo (InCor, HC-FMUSP), que tem como objetivo avaliar se pessoas fisicamente ativas podem apresentar maior proteção contra a COVID-19 e melhor recuperação após uma infecção. POR FAVOR, SÓ PARTICIPE DA PESQUISA SE VOCÊ CONTRAIU A COVID-19 E JÁ ESTÁ RECUPERADO DA DOENÇA.

Pesquisador responsável: Dr. Marcelo Rodrigues dos Santos
Contato:
Avaliação do risco da pesquisa: risco mínimo

Convite à participação: Convidamos o(a) senhor(a) para participar desta pesquisa voluntariamente a fim de aprimorarmos o conhecimento científico e futuramente melhorar a qualidade de vida de pacientes que tenham se infectado pela COVID-19.

Justificativa e os objetivos da pesquisa: Esse estudo científico do Instituto do Coração do Hospital das Clínicas da Universidade de São Paulo (InCor, HC-FMUSP), que tem como objetivo avaliar se pessoas fisicamente ativas podem apresentar maior proteção contra a COVID-19 e melhor recuperação após uma infecção.

Benefícios que poderão ser obtidos: Esperamos que esse estudo ajude a entender melhor o papel da atividade física e do exercício físico na proteção contra doenças virais. Além disso, poder colaborar com as políticas públicas de saúde em relação aos hábitos saudáveis de um estilo de vida mais ativo.

- O(a) senhor(a) terá acesso, a qualquer tempo, às informações sobre procedimentos, riscos e benefícios relacionados à pesquisa, inclusive para esclarecer eventuais dúvidas;
- O(a) senhor(a) terá liberdade de retirar seu consentimento a qualquer momento e de deixar de participar do estudo;
- Os resultados das suas informações serão guardados e não fornecidos a ninguém, garantindo a sua privacidade.
- Para mais informações, o(a) senhor(a) pode contatar o pesquisador responsável pelo

Esse estudo está aprovado pela Comissão de Ética em Pesquisa (CAPPesq nº 5044/20/073) e Plataforma Brasil (CAAE: 31958020.5.0000.0068). O estudo está registrado no ClinicalTrials (NCT04396353).

Eu li as informações acima sobre a minha decisão em participar desse estudo. Ficaram claros para mim os objetivos, os procedimentos, os potenciais desconfortos, os riscos e as garantias decorrentes do estudo. Concordo voluntariamente em participar deste estudo.
*Obrigatório

1. Eu aceito participar voluntariamente do estudo *

   *Marcar apenas uma oval.*

   ( ) Sim

2. Iniciais do seu nome *

   ______________________________

3. Seu e-mail *

   ______________________________

#### Sobre suas informações da doença, dados clínicos, antropométricos e sociodemográficos

A seção 2 tem um total de 20 perguntas (tempo total estimado para responder: 5 minutos)

4. 1) Você foi infectado pelo coronavírus (COVID-19)? *

   *Marcar apenas uma oval.*

   ( ) Sim
   ( ) Não

5. 2) Você teve o teste positivo confirmado por qual exame? *

*Marcar apenas uma oval.*

- RT-PCR (coleta feita com um bastão pelo nariz ou garganta)
- Exame de sangue (sorologia)
- Teste rápido (coleta de sangue na ponta do dedo)
- Não fiz nenhum teste
- Não sei dizer
- Outros

6. 3) Quais foram os sintomas que você teve durante a doença? Pode selecionar mais de uma resposta. *

*Marque todas que se aplicam.*

- [ ] Febre
- [ ] Dor no corpo (incluindo dores de cabeça e musculares)
- [ ] Falta de ar
- [ ] Tosse
- [ ] Perda do paladar
- [ ] Fadiga /cansaço
- [ ] Não tive sintomas

7. 4) Você necessitou ficar internado no hospital devido a COVID-19? *

*Marcar apenas uma oval.*

- Sim
- Não

8. 5) Se ficou internado, quantos dias você ficou hospitalizado? (Se não ficou internado, selecione o número "zero") *

*Marcar apenas uma oval.*

- 0
- 1
- 2
- 3
- 4
- 5
- 6
- 7
- 8
- 9
- 10
- 11
- 12
- 13
- 14
- 15
- 16
- 17
- 18
- 19
- 20
- 21
- 22
- 23
- 24
- 25
- 26
- 27
- 28
- 29
- 30

- 31
- Mais de 31

9. 6) Se ficou internado, você precisou de (pode selecionar mais de uma resposta): *

*Marque todas que se aplicam.*

- [ ] Respiração espontânea em ar ambiente
- [ ] Máscara de oxigênio
- [ ] Intubado para ajudar a respirar
- [ ] Não sei dizer
- [ ] Não fiquei internado

10. 7) Quais medicamentos você usou durante o tratamento? (Seja no hospital ou em casa. Pode selecionar mais de uma opção) *

*Marque todas que se aplicam.*

- [ ] Não tomei medicamentos
- [ ] Para dor ou febre (paracetamol, dipirona, ibuprofeno, outros)
- [ ] Antibióticos (amoxicilina, azitromicina, vancomicina, outros)
- [ ] Antivirais (lopinavir/ritonavir, oseltamivir, remdesivir)
- [ ] Hidroxicloroquina ou cloroquina
- [ ] Corticoides (prednisona, hidrocortisona, outros)
- [ ] Anticoagulante (heparina, outros)
- [ ] Outros
- [ ] Não sei dizer

11. 8) Você teve diagnóstico de pneumonia? *

*Marcar apenas uma oval.*

- Sim
- Não
- Não sei dizer

12. 9) Sexo *

*Marcar apenas uma oval.*

- Mulher
- Homem
- Mulher Transgênero
- Homem Transgênero

13. 10) Data de nascimento *

_Exemplo: 7 de janeiro de 2019_

14. 11) Idade (anos) *

15. 12) Peso (kg). (Exemplo: "70.5") *

16. 13) Altura (cm). (Exemplo: "170", sem pontos ou vírgula) *

17. 14) Você é fumante? *

*Marcar apenas uma oval.*

- Sim
- Não

18. 15) Etnia/Raça *

*Marcar apenas uma oval.*

- Negro
- Mulato
- Branco
- Asiático
- Outros

19. 16) Doenças pré-existentes. Pode selecionar mais de uma resposta. *

*Marque todas que se aplicam.*

- [ ] Diabetes
- [ ] Hipertensão
- [ ] Infarto
- [ ] Insuficiência cardíaca
- [ ] Doença arterial periférica
- [ ] Colesterol alto (hiperlipidemia)
- [ ] Obesidade
- [ ] Síndrome metabólica
- [ ] Doença pulmonar
- [ ] Doença renal
- [ ] Depressão
- [ ] Outros
- [ ] Nenhuma doença

20. 17) Escolaridade *

*Marcar apenas uma oval.*

- Ensino fundamental incompleto
- Ensino fundamental completo
- Ensino médio incompleto
- Ensino médio completo
- Superior completo (ou graduação)
- Pós-graduação
- Mestrado
- Doutorado
- Pós-Doutorado

21. 18) Renda familiar mensal (salário mínimo) *

*Marcar apenas uma oval.*

- Menos de 1
- 1 a 3
- 3 a 5
- 5 a 7
- 7 a 9
- Mais de 9

22. 19) País de residência *

______________________________

23. 20) Cidade de residência *

______________________________

#### Sobre seus hábitos de exercício físico (Questionário Internacional de Atividade Física - IPAQ)

***ATENÇÃO: VOCÊ DEVE RESPONDER AS PRÓXIMAS QUESTÕES COM RELAÇÃO AOS SEUS HÁBITOS DE ATIVIDADE FÍSICA ANTES DE TER FICADO DOENTE***

Instruções: Para responder as perguntas pense somente nas atividades que você realizava por pelo menos 10 minutos contínuos de cada vez *E ANTES DE FICAR DOENTE PELA COVID-19*

A seção 3 tem um total de 10 perguntas (tempo total estimado para responder: 5 minutos)

24. 1) De forma geral, ANTES DE FICAR DOENTE, você se considerava: *

*Marcar apenas uma oval.*

- Atleta
- Fisicamente ativo
- Sedentário

25. 2) ANTES DE FICAR DOENTE, quais atividades/execícios você praticava com regularidade (pode selecionar mais de uma opção): *

*Marque todas que se aplicam.*

- [ ] Caminhada
- [ ] Corrida
- [ ] Bicicleta (ciclismo, spinning, ergométrica)
- [ ] Musculação
- [ ] Crossfit/treinamento funcional
- [ ] Esportes coletivos (futebol, vôlei, basquete, etc.)
- [ ] Esportes individuais (tênis, golfe, escalada, etc.)
- [ ] Natação
- [ ] Yoga
- [ ] Pilates
- [ ] Outros
- [ ] Não pratico nenhuma atividade/exercício

26. 3A) ANTES DE FICAR DOENTE, você CAMINHAVA por pelo menos 10 minutos contínuos em casa ou no trabalho, como forma de transporte para ir de um lugar para outro, por lazer, por prazer ou como forma de exercício? *

*Marcar apenas uma oval.*

- 1 dia na semana
- 2 dias na semana
- 3 dias na semana
- 4 dias na semana
- 5 dias na semana
- 6 dias na semana
- 7 dias na semana
- Nenhum

27. 3B) Nos dias em que você faz CAMINHADA por pelo menos 10 minutos contínuos, quanto tempo no total você gastava caminhando por dia? (Se marcou "Nenhum" na pergunta acima, selecione "0:00") *

*Marcar apenas uma oval.*

- 0:00
- 10 minutos
- 15 minutos
- 20 minutos
- 25 minutos
- 30 minutos
- 35 minutos
- 40 minutos
- 45 minutos
- 50 minutos
- 55 minutos
- 60 minutos
- 1:05 hora
- 1:10 hora
- 1:15 hora
- 1:20 hora
- 1:25 hora
- 1:30 hora
- 1:35 hora
- 1:40 hora
- 1:45 hora
- 1:50 hora
- 1:55 hora
- 2:00 horas
- 2:05 horas
- 2:10 horas
- 2:15 horas
- 2:20 horas
- 2:25 horas
- 2:30 horas

- 2:35 horas
- 2:40 horas
- 2:45 horas
- 2:50 horas
- 2:55 horas
- 3:00 horas ou mais

28. 4A) ANTES DE FICAR DOENTE, você realizava atividades MODERADAS por pelo menos 10 minutos contínuos, como por exemplo: pedalar leve na bicicleta, nadar, dançar, fazer ginástica aeróbica leve, jogar vôlei recreativo, carregar pesos leves, fazer serviços domésticos na casa, no quintal ou no jardim como varrer, aspirar, cuidar do jardim, ou qualquer atividade que fez aumentar moderadamente sua respiração ou batimentos do coração (POR FAVOR NÃO INCLUA CAMINHADA) *

*Marcar apenas uma oval.*

- 1 dia na semana
- 2 dias na semana
- 3 dias na semana
- 4 dias na semana
- 5 dias na semana
- 6 dias na semana
- 7 dias na semana
- Nenhum

29. 4B) Nos dias em que você faz atividades MODERADAS por pelo menos 10 minutos contínuos, quanto tempo no total você gastava fazendo essas atividades por dia? (Se marcou "Nenhum" na pergunta acima, selecione "0:00") *

*Marcar apenas uma oval.*

- 0:00
- 10 minutos
- 15 minutos
- 20 minutos
- 25 minutos
- 30 minutos
- 35 minutos
- 40 minutos
- 45 minutos
- 50 minutos
- 55 minutos
- 60 minutos
- 1:05 hora
- 1:10 hora
- 1:15 hora
- 1:20 hora
- 1:25 hora
- 1:30 hora
- 1:35 hora
- 1:40 hora
- 1:45 hora
- 1:50 hora
- 1:55 hora
- 2:00 horas
- 2:05 horas
- 2:10 horas
- 2:15 horas
- 2:20 horas
- 2:25 horas
- 2:30 horas

- 2:35 horas
- 2:40 horas
- 2:45 horas
- 2:50 horas
- 2:55 horas
- 3:00 horas ou mais

30. 5A) ANTES DE FICAR DOENTE, você realizava atividades VIGOROSAS por pelo menos 10 minutos contínuos, como por exemplo: correr, fazer ginástica aeróbica, jogar futebol, pedalar rápido na bicicleta, jogar basquete, fazer serviços domésticos pesados em casa, no quintal ou cavoucar no jardim, carregar pesos elevados ou qualquer atividade que fez aumentar MUITO sua respiração ou batimentos do coração? *

*Marcar apenas uma oval.*

- 1 dia na semana
- 2 dias na semana
- 3 dias na semana
- 4 dias na semana
- 5 dias na semana
- 6 dias na semana
- 7 dias na semana
- Nenhum

31. 5B) Nos dias em que você faz essas atividades VIGOROSAS por pelo menos 10 minutos contínuos, quanto tempo no total você gastava fazendo essas atividades por dia? (Se marcou "Nenhum" na pergunta acima, selecione "0:00") *

*Marcar apenas uma oval.*

- 0:00
- 10 minutos
- 15 minutos
- 20 minutos
- 25 minutos
- 30 minutos
- 35 minutos
- 40 minutos
- 45 minutos
- 50 minutos
- 55 minutos
- 60 minutos
- 1:05 hora
- 1:10 hora
- 1:15 hora
- 1:20 hora
- 1:25 hora
- 1:30 hora
- 1:35 hora
- 1:40 hora
- 1:45 hora
- 1:50 hora
- 1:55 hora
- 2:00 horas
- 2:05 horas
- 2:10 horas
- 2:15 horas
- 2:20 horas
- 2:25 horas
- 2:30 horas

- 2:35 horas
- 2:40 horas
- 2:45 horas
- 2:50 horas
- 2:55 horas
- 3:00 horas ou mais

32. 6) Quanto tempo no total, ANTES DE FICAR DOENTE, você gastava sentado durante um dia na SEMANA? Assistindo TV, dirigindo, trabalhando. *

*Marcar apenas uma oval.*

- 0:00
- 10 minutos
- 20 minutos
- 30 minutos
- 40 minutos
- 50 minutos
- 60 minutos
- 1:30 hora
- 2:00 horas
- 2:30 horas
- 3:00 horas
- 3:30 horas
- 4:00 horas
- 4:30 horas
- 5:00 horas
- 5:30 horas
- 6:00 horas
- 6:30 horas
- 7:00 horas
- 7:30 horas
- 8:00 horas
- 8:30 horas
- 9:00 horas
- 9:30 horas
- 10:00 horas
- 10:30 horas
- 11:00 horas
- 11:30 horas
- 12:00 horas ou mais

33. 7) Quanto tempo no total, ANTES DE FICAR DOENTE, você gastava sentado durante um dia de FINAL DE SEMANA? Assistindo TV, dirigindo, trabalhando. *

*Marcar apenas uma oval.*

- 0:00
- 10 minutos
- 20 minutos
- 30 minutos
- 40 minutos
- 50 minutos
- 60 minutos
- 1:30 hora
- 2:00 horas
- 2:30 horas
- 3:00 horas
- 3:30 horas
- 4:00 horas
- 4:30 horas
- 5:00 horas
- 5:30 horas
- 6:00 horas
- 6:30 horas
- 7:00 horas
- 7:30 horas
- 8:00 horas
- 8:30 horas
- 9:00 horas
- 9:30 horas
- 10:00 horas
- 10:30 horas
- 11:00 horas
- 11:30 horas
- 12:00 horas ou mais

Este conteúdo não foi criado nem aprovado pelo Google.

Google Formulários
