## Supplementary material for "Physical Activity Decreases the Prevalence of COVID-19-associated Hospitalization: Brazil EXTRA Study": English Google Form

### Title: The Impact of EXercise TRAining, Physical Activity and Sedentary Lifestyle on Clinical Outcomes in Surviving Patients Infected with the SARS-CoV-2 Virus (EXTRA SARS-CoV-2 Study)

HOSPITAL DAS CLÍNICAS DA FACULDADE DE MEDICINA DA UNIVERSIDADE DE SÃO PAULO-HCFMUSP

INFORMED CONSENT FORM

This form will be used for a scientific study by the Instituto do Coração of the Hospital das Clínicas of the University of São Paulo (InCor, HC-FMUSP), Brazil, which aims to assess whether physically active people can have greater protection against COVID-19 and better recovery after an infection. PLEASE ONLY PARTICIPATE IN THE RESEARCH IF YOU HAVE CONTRACTED COVID-19 AND ARE FULLY RECOVERED FROM THE DISEASE.

Principal investigator: Dr. Marcelo Rodrigues dos Santos
Contact:
Research risk assessment: minimal risk

Invitation to participate: We invite you to participate in this research voluntarily in order to improve scientific knowledge and improve the quality of life of patients who have become infected with COVID-19.

Justification and objectives of the research: This scientific study aims to assess whether physically active people can have greater protection against COVID-19 and better recovery after an infection.

Benefits that can be obtained: We hope that this study will help to better understand the role of physical activity and exercise training in protecting against viral diseases. In addition, collaborate with public health policies in relation to the healthy habits of a more active lifestyle.

- You will have access, at any time, to information on procedures, risks, and benefits related to research, including to clarify any doubts;
- You will be free to withdraw your consent at any time and to stop participating in the study;
- The results of your information will be saved and not provided to anyone, guaranteeing your privacy.
- For more information, you can contact the principal investigator by

This study is approved by the Research Ethics Committee (CAPPesq nº 5044/20/073) and Plataforma Brasil (CAAE: 31958020.5.0000.0068). The study is registered on ClinicalTrials (NCT04396353).

I read the information above about my decision to participate in this study. The objectives, procedures, potential discomforts, risks, and guarantees resulting from the study were clear to me. I voluntarily agree to participate in this study.
*Obrigatório

1. I accept to voluntarily participate in the study *

*Marcar apenas uma oval.*

- Yes

2. Your name initials *

3. Your e-mail *

#### About your disease information, clinical, anthropometric, and sociodemographic data

Section 2 has a total of 20 questions (total estimated time to answer: 5 minutes)

4. 1) Have you been infected with the coronavirus (COVID-19)? *

*Marcar apenas uma oval.*

- Yes
- No

5. 2) Which test did you take to confirm the disease? *

*Marcar apenas uma oval.*

- RT-PCR (collection made with a stick through the nose or throat)
- Blood test (serology)
- Rapid test (blood collection at the fingertip)
- I did not do any tests
- I do not know
- Others

6. 3) What symptoms did you have during the illness? You can select more than one answer. *

*Marque todas que se aplicam.*

- [ ] Fever
- [ ] Body pain (including headaches and muscle aches)
- [ ] Shortness of breathe
- [ ] Cough
- [ ] Loss of taste
- [ ] Fatigue/Tiredness
- [ ] I had no symptoms

7. 4) Did you need to be admitted to the hospital due to COVID-19? *

*Marcar apenas uma oval.*

- Yes
- No

8. 5) If you were hospitalized, how many days were you hospitalized? (If you have not been admitted, select the number "zero") *

*Marcar apenas uma oval.*

- 0
- 1
- 2
- 3
- 4
- 5
- 6
- 7
- 8
- 9
- 10
- 11
- 12
- 13
- 14
- 15
- 16
- 17
- 18
- 19
- 20
- 21
- 22
- 23
- 24
- 25
- 26
- 28
- 29
- 30
- 31

- More than 31

9. 6) If you were hospitalized, you needed (you can select more than one answer): *

*Marque todas que se aplicam.*

- [ ] Spontaneous breathing in room air
- [ ] Oxygen mask
- [ ] Intubated to help breathe
- [ ] I do not know
- [ ] I have not been hospitalized

10. 7) What medications did you need during treatment? (Whether at the hospital or at home. You can select more than one option)

*Marque todas que se aplicam.*

- [ ] I did not take any medications
- [ ] For pain or fever (paracetamol, dipyrone, ibuprofen, others)
- [ ] Antibiotics (amoxicillin, azithromycin, vancomycin, others)
- [ ] Antivirals (lopinavir/ritonavir, oseltamivir, remdesivir)
- [ ] Hydroxychloroquine or chloroquine
- [ ] Corticosteroids (prednisone, hydrocortisone, others)
- [ ] Anticoagulant (heparin, others)
- [ ] Others
- [ ] I do not know

11. 8) Have you been diagnosed with pneumonia? *

*Marcar apenas uma oval.*

- Yes
- No
- I do not know

12. 9) Sex *

*Marcar apenas uma oval.*

- Female
- Male
- Transgender Female
- Transgender Male

13. 10) Date of birth (day/month/year) *

*Exemplo: 7 de janeiro de 2019*

14. 11) Age (years) *

15. 12) Weight (kg). (Example: 70.5) *

16. 13) Height (cm). (Example: "170", without dot or comma) *

17. 14) Do you smoke? *

*Marcar apenas uma oval.*

- Yes
- No

18. 15) Ethnicity/Race *

*Marcar apenas uma oval.*

- Black
- Mulatto
- White
- Asian
- Hispanic
- Others

19. 16) Pre-existing diseases. You can select more than one answer. *

*Marque todas que se aplicam.*

- [ ] Diabetes
- [ ] Hipertension
- [ ] Myocardial infarction
- [ ] Heart failure
- [ ] Peripheral arterial disease
- [ ] High cholesterol (hyperlipidemia)
- [ ] Obesity
- [ ] Metabolic syndrome
- [ ] Pulmonary disease
- [ ] Kidney disease
- [ ] Depression
- [ ] Others
- [ ] No disease

20. 17) Education *

*Marcar apenas uma oval.*

- Elementary school
- Middle school
- High school
- College
- Postgraduate
- Master
- Doctorate
- Postdoctoral

21. 18) Family income (minimum wage) *

*Marcar apenas uma oval.*

- Less than 1
- 1 to 3
- 3 to 5
- 5 to 7
- 7 to 9
- More than 9

22. 19) Country of residence *

_______________

23. 20) City of residence *

_______________

#### About your physical exercise habits (International Physical Activity Questionnaire - IPAQ)

*** PLEASE NOTE: YOU SHOULD ANSWER YOUR NEXT QUESTIONS REGARDING YOUR PHYSICAL ACTIVITY HABITS BEFORE YOU GET SICK ***

Instructions: To answer the questions, think only of the activities you performed for at least 10 continuous minutes at a time *AND BEFORE YOU GET SICK WITH COVID-19*

Section 3 has a total of 10 questions (total estimated time to answer: 5 minutes)

24. 1) In general, BEFORE YOU GET SICK, you considered yourself: *

*Marcar apenas uma oval.*

- Athlete
- Physically active
- Sedentary

25. 2) BEFORE YOU GET SICK, what activities/exercises did you regularly practice? (you can select more than one option): *

*Marque todas que se aplicam.*

- [ ] Walking
- [ ] Running
- [ ] Bicycle (cycling, spinning, ergometric)
- [ ] Strength training (weight training, bodybuilding)
- [ ] Crossfit/functional training
- [ ] Team sports (football, volleyball, basketball, etc.)
- [ ] Individual sports (tennis, golf, climbing, etc.)
- [ ] Swimming
- [ ] Yoga
- [ ] Pilates
- [ ] Others
- [ ] I do not practice any activity/exercise

26. 3A) BEFORE YOU GET SICK, did you WALK for at least 10 continuous minutes at home or at work, as a form of transportation to get from place to place, for leisure, for pleasure, or as a form of exercise? *

*Marcar apenas uma oval.*

- 1 day a week
- 2 days a week
- 3 days a week
- 4 days a week
- 5 days a week
- 6 days a week
- 7 days a week
- None

27. 3B) On days when you walked for at least 10 continuous minutes, how much time in total did you spend walking each day? (If you checked "None" in the question above, select "0:00") *

*Marcar apenas uma oval.*

- 0:00
- 10 minutes
- 15 minutes
- 20 minutes
- 25 minutes
- 30 minutes
- 35 minutes
- 40 minutes
- 45 minutes
- 50 minutes
- 55 minutes
- 60 minutes
- 1:05 hour
- 1:10 hour
- 1:15 hour
- 1:20 hour
- 1:25 hour
- 1:30 hour
- 1:35 hour
- 1:40 hour
- 1:45 hour
- 1:50 hour
- 1:55 hour
- 2:00 hours
- 2:05 hours
- 2:10 hours
- 2:15 hours
- 2:20 hours
- 2:25 hours
- 2:30 hours

- 2:35 hours
- 2:40 hours
- 2:45 hours
- 2:50 hours
- 2:55 hours
- 3:00 hours or more

28. 4A) BEFORE YOU GET SICK, have you performed MODERATE activities for at least 10 continuous minutes, such as: cycling lightly on the bike, swimming, dancing, doing light aerobics, playing recreational volleyball, carrying light weights, doing housework at home, in the backyard or in the garden like sweeping, vacuuming, gardening, or any activity that moderately increased your breathing or heartbeat (PLEASE DO NOT INCLUDE WALK) *

*Marcar apenas uma oval.*

- 1 day a week
- 2 days a week
- 3 days a week
- 4 days a week
- 5 days a week
- 6 days a week
- 7 days a week
- None

29. 4B) On the days that you did these MODERATE activities for at least 10 continuous minutes, how much time in total did you spend doing these activities each day? (If you checked "None" in the question above, select "0:00") *

*Marcar apenas uma oval.*

- 0:00
- 10 minutes
- 15 minutes
- 20 minutes
- 25 minutes
- 30 minutes
- 35 minutes
- 40 minutes
- 45 minutes
- 50 minutes
- 55 minutes
- 60 minutes
- 1:05 hour
- 1:10 hour
- 1:15 hour
- 1:20 hour
- 1:25 hour
- 1:30 hour
- 1:35 hour
- 1:40 hour
- 1:45 hour
- 1:50 hour
- 1:55 hour
- 2:00 hours
- 2:05 hours
- 2:10 hours
- 2:15 hours
- 2:20 hours
- 2:25 hours

- 2:30 hours
- 2:35 hours
- 2:40 hours
- 2:45 hours
- 2:50 hours
- 2:55 hours
- 3:00 hours or more

30. 5A) BEFORE YOU GET SICK, have you performed VIGOROUS activities for at least 10 continuous minutes, such as running, doing aerobics, playing football, cycling fast, playing basketball, doing heavy housework at home, in the yard or digging in the garden, carrying heavy weights or any activity that has greatly increased your breathing or heart rate? *

*Marcar apenas uma oval.*

- 1 day a week
- 2 days a week
- 3 days a week
- 4 days a week
- 5 days a week
- 6 days a week
- 7 days a week
- None

31. 5B) On the days that you did these VIGOROUS activities for at least 10 continuous minutes, how much time in total did you spend doing these activities each day? (If you checked "None" in the question above, select "0:00") *

*Marcar apenas uma oval.*

- 0:00
- 10 minutes
- 15 minutes
- 20 minutes
- 25 minutes
- 30 minutes
- 35 minutes
- 40 minutes
- 45 minutes
- 50 minutes
- 55 minutes
- 60 minutes
- 1:05 hour
- 1:10 hour
- 1:15 hour
- 1:20 hour
- 1:25 hour
- 1:30 hour
- 1:35 hour
- 1:40 hour
- 1:45 hour
- 1:50 hour
- 1:55 hour
- 2:00 hours
- 2:05 hours
- 2:10 hours
- 2:15 hours
- 2:20 hours
- 2:25 hours

- 2:30 hours
- 2:35 hours
- 2:40 hours
- 2:45 hours
- 2:50 hours
- 2:55 hours
- 3:00 hours or more

32. 6) How much time in total, BEFORE YOU GET SICK, do you spend sitting on a WEEKDAY? Watching TV, driving, working. *

*Marcar apenas uma oval.*

- 0:00
- 10 minutes
- 20 minutes
- 30 minutes
- 40 minutes
- 50 minutes
- 60 minutes
- 1:30 hour
- 2:00 hours
- 2:30 hours
- 3:00 hours
- 3:30 hours
- 4:00 hours
- 4:30 hours
- 5:00 hours
- 5:30 hours
- 6:00 hours
- 6:30 hours
- 7:00 hours
- 7:30 hours
- 8:00 hours
- 8:30 hours
- 9:00 hours
- 9:30 hours
- 10:00 hours
- 10:30 hours
- 11:00 hours
- 11:30 hours
- 12:00 hours or more

33. 7) How much time in total, BEFORE YOU GET SICK, do you spend sitting on a WEEKEND day? Watching TV, driving, working. *

*Marcar apenas uma oval.*

- 0:00
- 10 minutes
- 20 minutes
- 30 minutes
- 40 minutes
- 50 minutes
- 60 minutes
- 1:30 hour
- 2:00 hours
- 2:30 hours
- 3:00 hours
- 3:30 hours
- 4:00 hours
- 4:30 hours
- 5:00 hours
- 5:30 hours
- 6:00 hours
- 6:30 hours
- 7:00 hours
- 7:30 hours
- 8:00 hours
- 8:30 hours
- 9:00 hours
- 9:30 hours
- 10:00 hours
- 10:30 hours
- 11:00 hours
- 11:30 hours
- 12:00 hours or more

Este conteúdo não foi criado nem aprovado pelo Google.

Google Formulários
